# New Diagnostic Methods for *Escherichia marmotae* and the First Report of its Identification in Clinical Isolates in North America

**DOI:** 10.1101/2025.07.10.25331261

**Authors:** Pelumi M. Oladipo, Robert J. Tibbetts, Audun Sivertsen, Justin M. Barger, Torbjørn S. Bruvold, Alemu Fite, Matthew Sims, Marcus Zervos, Ali Jomaa, Jeffrey L. Ram

## Abstract

Genomic sequences of *E. marmotae* and *E. coli* differ by 10%. Discovered as an environmental “cryptic clade” of *Escherichia*, *E. marmotae* also occurs in human infections. Microbiological and MALDI-TOF-MS methods frequently misidentify *E. marmotae* as *E. coli.* Our goal was to develop methods that reliably distinguish *E. marmotae* from *E. coli* to improve therapeutic decisions and treatments.

A Taqman PCR method was developed to distinguish *E. marmotae* from *E. coli* based on sequences of *uidA* and *uidB* and a positive control targeting *adk*. *uidA-* and *uidB* species-specific PCR amplified DNA from *E. marmotae* with 100% specificity, and not from *E. coli* or other *Escherichia* species.

The Biomérieux VITEK MALDI-TOF-MS consistently misidentified *E. marmotae* as *E. coli*, with median IVD confidence scores for both *E. marmotae* and *E. coli* of 99.9%; however, RUO scores for *E. marmotae* (median 0%) were significantly lower (P<0.0001) than for *E. coli* (median=87.4%). The spectral peak between m/z 7250 to 7280 consistently occurred between 7260 and 7268 in *E. marmotae* and only between 7268 and 7280 in *E. coli*, with no overlap (p<0.001).

Application of these spectral criteria to 176 clinical isolates revealed the first isolate of *E. marmotae* from a human infection in North America. The isolate had originally been diagnosed as *E. coli* based on a 99.1% IVD confidence score. This first North American clinical isolate was confirmed as *E. marmotae* by Taqman-PCR and whole genome sequencing. This isolate had numerous antibiotic resistance gene markers and unlike most clinical *E. coli*, this *E. marmotae* isolate lacked motility at 37 °C.

Clinical tests based on these methods of differentiating *E. marmotae* and *E. coli* may assist in therapeutic decisions. Moreover, discovering the identity of the structure underlying the shift in the MALDI-TOF-MS peak may help determine if it has a special role in infection.

## INTRODUCTION

*Escherichia marmotae* was originally discovered as a “cryptic clade” of *Escherichia*, having identical metabolic and colony morphology profiles on standard microbiological tests but having an average pairwise difference from *E. coli* over its whole genome of about 10% (1,2). Although originally discovered in animal feces (e.g., marmots (3), racoons (4), and birds (2) and environmental samples (2), *E. marmotae* has now been identified in clinical cases as serious as those caused by *E. coli.* Clinical isolates of *E. marmotae* have been identified from human cases of septicemia (5), urinary tract infection (6), and thoracic spondylodiscitis, pyelonephritis, acute sepsis of unknown origin, and postoperative sepsis (7). However, within these reports, *E. marmotae* were always initially misclassified as *E. coli* by routine clinical tests. Techniques that distinguish these two species are often not applied to clinical isolates once the tentative diagnosis as *E. coli* has been made. Therefore, we have only limited knowledge about the prevalence of *E. marmotae* in human infections, its relative risk of causing serious disease, and whether different treatments than are currently used for *E. coli* infections may be more effectively used on *E. marmotae* infections. Rapid, convenient methods of distinguishing *E. marmotae* from *E. coli* clinically are needed to determine its prevalence and to guide its treatment.

The identification of *E. marmotae* by routine clinical methods has proven challenging. One previous approach to identifying *E. marmotae* has been to use a set of PCR amplifications followed by agarose gel electrophoresis to identify PCR product sizes (5). Application of this method to 1081 strains from septicemic patients that were originally classified as *E. coli* during the COLIBAFI study in France (8) discovered that two were actually *E. marmotae* (5).

Matrix-Assisted Laser Desorption Ionization-Time of Flight Mass Spectrometry (MALDI-TOF MS) has become a frequently used tool for identifying bacterial isolates (9); however, its use for identification of *E. marmotae* has been problematic as the software for a long time contained no *E. marmotae* spectra. The first *E. marmotae* spectrum in the Bruker database was included in 2021 (database version L2020 9607MSP), but did not reliably distinguish *E. marmotae* from *E. coli*, rarely achieving an identity score >2.0, as required for the most confident identification in the Bruker system. Later versions have included additional spectra that align better with typical *E. marmotae,* often achieving identity scores >2.2 (Sivertsen et al., 2022). In another laboratory (6), an isolate was identified as *E. marmotae* by 16S RNA gene sequencing was initially classified as *E. coli* with 99% probability by the colorimetric and substrate-specific tests with the VITEK 2 XL GNI ID card system (BioMérieux, Australia). A subsequent test on a Bruker MALDI-TOF-MS using a database that included *E. marmotae* reported a confidence score of 2.39 for *E. marmotae* (6).

In this paper, we describe two methods, potentially applicable in a clinical laboratory, for reliably distinguishing *E. marmotae* from *E. coli.* The first is a Taqman PCR method based on sequence differences between *E. marmotae* and *E. coli*. In the second method, we identify reliable spectral differences between *E. marmotae* and *E. coli* on a Biomerieux VITEK MALDI-TOF-MS system, as all previous studies have reported analysis of *E. marmotae* using the Bruker system. In preliminary studies to the results shown in this paper, we found that these VITEK systems classify most isolates of *E. marmotae* as *E. coli* (i.e., they report In Vitro Diagnostic (IVD) confidence scores for *E. coli* greater than 99%) or, for a minority of *E. marmotae* isolates, fail to identify them at all. In this paper, we identify a specific peak in the MALDI-TOF-MS spectra that is exclusively and reliably associated with *E. marmotae* that enabled the discovery of the first *E. marmotae* strain isolated in North America.

## MATERIALS AND METHODS

### Bacterial Isolates

The Ram laboratory previously isolated six strains of *E. marmotae* from aquatic environments and raccoons (4) identified by whole genome sequencing. These isolates were archived at −80 ± 2°C in glycerol stocks (in Colilert 18 (IDEXX US) media with 15% glycerol) and have reliably yielded viable subcultures. Seventeen additional *E. marmotae* isolates, designated with the “TW” prefix, were retrieved from the Thomas Whittam strain collection at Michigan State University. Some of the TW strains were originally derived from the same *E. marmotae* isolates produced by the Ram lab and other environmental sources (2). Therefore, comparisons between Ram lab and TW strains serve as internal replicates, enabling assessment of reproducibility in measurements from independently maintained stocks of the same strain.

In addition to these isolates, the present study also investigated other strains i) *E. marmotae* isolated by others from human clinical sources (Sivertsen et al., 2022), and additional isolates of *E. marmotae*: ii) Representative isolates of other cryptic clades and *Escherichia* species from the microbial archives of Michigan State University, described originally by (2,10), and iii) 175 strains of putative *E. coli* that were originally identified as such by MALDI-TOF-MS at Henry Ford Health.

Table 1 shows the metadata of the strains of *E. marmotae* and *E.coli* that are analyzed in the present study, except for the strains from Henry Ford Health, which are described in further detail in the methods section on MALDI-TOF-MS.

**Table 1.** Sample labels (Sample ID), species, sources of isolates, and previous publications (if any) in which the isolate was previously reported or used.

| Sample ID | Accession Numbers | Species Identification | Source |
| --- | --- | --- | --- |
| TW14263 alias Ram 3024 | *JBNVMX000000000* | *Escherichia marmotae* | Raccoon Feces<sup>a,b</sup> |
| TW14264 alias Ram 3032 | *JBNVMU000000000* | *Escherichia marmotae* | Environment<sup>a,b</sup> |
| TW14265 alias Ram 3050 | *JBNVMV000000000* | *Escherichia marmotae* | Environment<sup>a,b</sup> |
| TW14266 alias Ram 3054 | *JBNVMW000000000* | *Escherichia marmotae* | Environment<sup>a,b</sup> |
| TW14267 alias Ram 3318 | *JBNVMY000000000* | *Escherichia marmotae* | Environment<sup>a,b</sup> |
| TW 09308 | *NZ AEME01000001* | *Escherichia marmotae* | Environment<sup>a,b</sup> |
| TW 14351 | NA | *Escherichia marmotae* | Dog<sup>b</sup> |
| TW 15825 | NA | *Escherichia marmotae* | Bird<sup>b</sup> |
| TW 15841 | NA | *Escherichia marmotae* | Environment<sup>b</sup> |
| TW 15835 | NA | *Escherichia marmotae* | Environment<sup>b</sup> |
| TW 15848 | NA | *Escherichia marmotae* | Mammal<sup>b</sup> |
| TW 15839 | NA | *Escherichia marmotae* | Environment<sup>b</sup> |
| TW 15833 | NA | *Escherichia marmotae* | Bird<sup>b</sup> |
| TW 15846 | NA | *Escherichia marmotae* | Mammal<sup>b</sup> |
| TW 15836 | NA | *Escherichia marmotae* | Environment<sup>b</sup> |
| TW 15840 | NA | *Escherichia marmotae* | Environment<sup>b</sup> |
| TW 15834 | NA | *Escherichia marmotae* | Environment<sup>b</sup> |
| C4B | NA | *Escherichia marmotae* | Blood<sup>c</sup> |
| C5 | NA | *Escherichia marmotae* | Blood<sup>c</sup> |
| C6 | NA | *Escherichia marmotae* | Spondylodiscitis<sup>c</sup> |
| C1 | CAKAEK000000000 | *Escherichia marmotae* | Blood<sup>c</sup> |
| C7 | NA | *Escherichia marmotae* | Blood<sup>c</sup> |
| C8 | NA | *Escherichia marmotae* | Blood<sup>c</sup> |
| AS8 | NA | *Escherichia marmotae* | Blood<sup>c</sup> |
| C9 | NA | *Escherichia marmotae* | Urine<sup>c</sup> |
| C10 | NA | *Escherichia marmotae* | Urine<sup>c</sup> |
| C11 | NA | *Escherichia marmotae* | Blood<sup>c</sup> |
| C12 | NA | *Escherichia marmotae* | Urine<sup>c</sup> |
| C3 | *CAKAEJ000000000* | *Escherichia marmotae* | Blood<sup>c</sup> |
| AS14 | NA | *Escherichia marmotae* | Urine<sup>c</sup> |
| AS15 | NA | *Escherichia marmotae* | Urine<sup>c</sup> |
| AS16 | NA | *Escherichia marmotae* | Urine<sup>c</sup> |
| AS17 | NA | *Escherichia marmotae* | Urine<sup>c</sup> |
| AS18 | NA | *Escherichia marmotae* | Pus<sup>c</sup> |
| C2 | *CAKAEI000000000* | *Escherichia marmotae* | Blood<sup>c</sup> |
| C4A | CAKAEL000000000 | *Escherichia marmotae* | Permanent Urine Catheter<sup>c</sup> |
| AS21 | NA | *Escherichia marmotae* | Urine<sup>c</sup> |
| AS22 | NA | *Escherichia marmotae* | Recently changed Urine Catheter<sup>c</sup> |
| AS23 | NA | *Escherichia marmotae* | Blood<sup>c</sup> |
| TW 15976 | NA | *Escherichia cryptic* Clade 1 | Not reported<sup>d</sup> |
| TW 15838 | *AEJX01000001* | *Escherichia cryptic* Clade 1 | Not reported<sup>d</sup> |
| TW 15951 | NA | *Escherichia cryptic* Clade 1 | Not reported<sup>d</sup> |
| TW 15832 | NA | *Escherichia whittamii* | Not reported<sup>d</sup> |
| TW 09254 | NA | *Escherichia ruyisiae* | Freshwater beach<sup>b</sup> |
| TW 09231 | NA | *Escherichia ruyisiae* | Freshwater beach<sup>b</sup> |
| TW 14182 | *AEJZ01000533* | *Escherichia ruyisiae* | Freshwater beach<sup>b</sup> |
| TW 11588 | *AEMF01000001* | *Escherichia ruyisiae* | Soil<sup>b</sup> |
| 250 | NA | *Escherichia coli* | Environment<sup>e</sup> |
| 430 | NA | *Escherichia coli* | Environment<sup>e</sup> |
| 1406 | NA | *Escherichia coli* | Environment<sup>e</sup> |
| 970 | NA | *Escherichia coli* | Environment<sup>e</sup> |

<sup>a</sup>(4); <sup>b</sup>(2); <sup>c</sup>(Sivertsen et al., 2022) or directly provided by Sivertsen; <sup>d</sup>(16); <sup>e</sup>(12) NA-Not Available

### Primers and TaqMan probe design

Sequence data of Ram lab strains in another study in our lab (11) were used to find suitable assay targets. The genes *uidA* and *uidB*, which encode for beta-glucuronidase and the glucuronide carrier protein, respectively, and have both conserved and variable regions among various *Escherichia* species, were chosen for designing primers and Taqman probes for detecting *E. marmotae* and *E. coli*. Studies of the variability of sequences of *uidA* among environmental and fecal isolates (4,12) had previously led to the discovery of phenotypically similar strains of *E. coli* and the strain subsequently identified as *E. marmotae*. The genes exhibit >8% mismatches in nucleotide identity between *E. marmotae* and *E.coli*. Geneious Prime software (version 2024.0, Dotmatics) was used to design forward and reverse primers, and a TaqMan probe targeting the mismatch region specific to *E. marmotae*, following best practices for specificity and efficiency in amplification (13,14).

We also tested *E.coli* specific *lipB* primers, as previously described by (15) across all 176 clinical isolates.In addition, we used universal primers targeting the adenylate kinase (*adk*) gene (2) and designed a probe conserved in all *Escherichia* species as a normalization standard, enabling amplification of both *E. marmotae* and *E.coli*. Sequences of the primers and probes are provided in Table 2. Specificity of the *E. marmotae*-primers and probes in Table 2 was studied by *in silico* analysis using PubMLST, to compare them against *E. coli* strains and other *Escherichia* genomes available in GenBank. Primers and probes were synthesized by Integrated DNA Technologies (IDT).

**Table 2:**
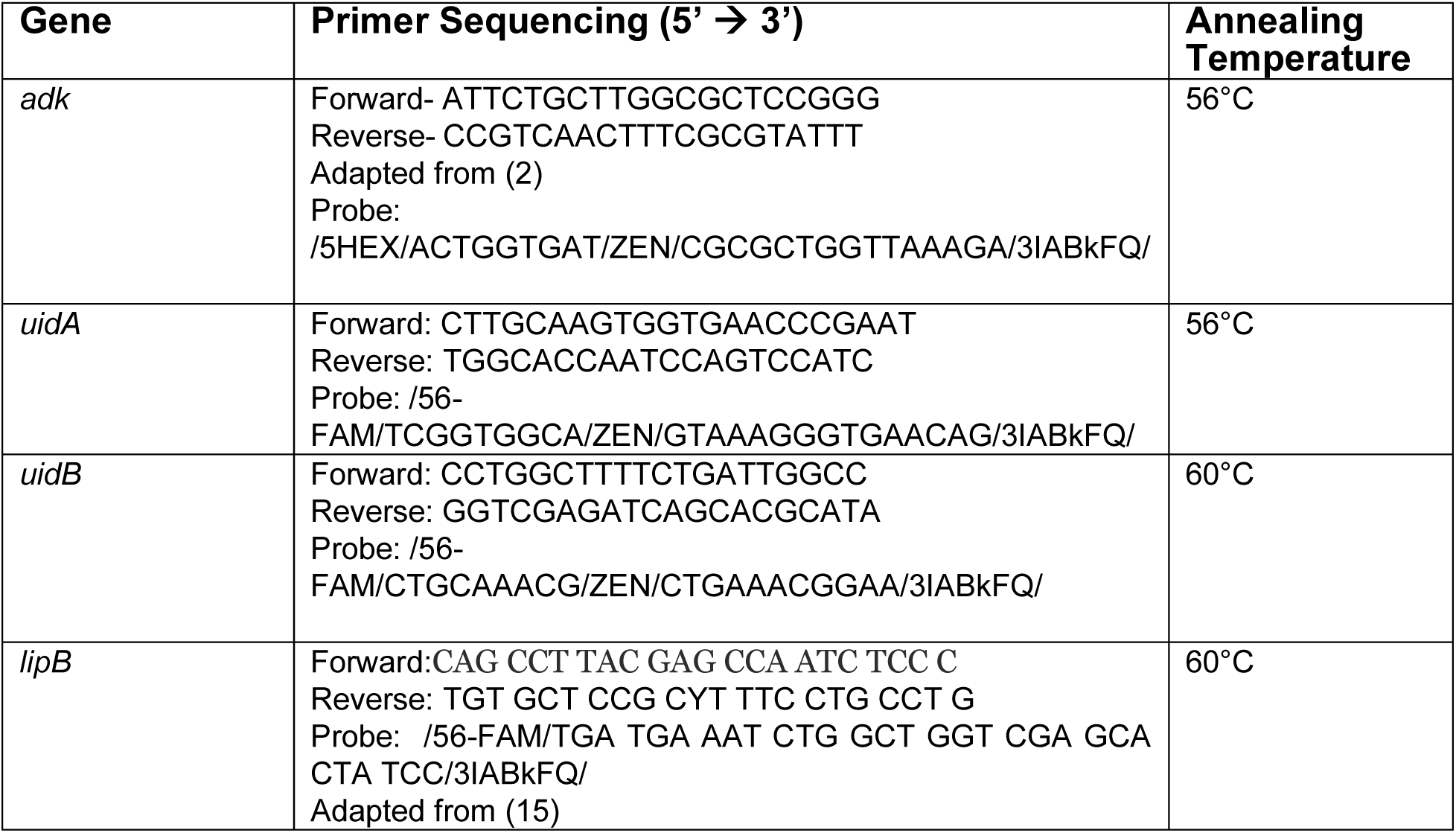
Primers and probes used in the analysis.

### Quantitative PCR Procedure

Single colonies were picked using 200 µL pipet tips and suspended in 300 µL of nuclease-free water. The qPCR was performed on a Bio-Rad CFX 96 Touch Real-time PCR detection system with Bio-Rad CFX manager software version 3.1. The optimal annealing temperature was determined by performing a temperature gradient (55° - 65°C) within a single run using DNA samples of *E. marmotae* and *E.coli*. The optimized cycling parameters were 95°C for 5 min; 30 cycles of 95°C for 30 s, annealing temperature as described in table 2, and 72°C for 40 s; then hold at 4°C. The amplification was performed in duplicate in 22 µL, containing 5.5 µL of SsoAdvanced Universal Probes Supermix (Bio-Rad), 3.3 µL each of *uidA, uidB* or *adk* probe and primers stock solution (250 nM final concentration for each probe and 900 nM final concentration for each primer), 1 µL of sample, and nuclease free water to a total volume of 22 µL. The reaction was run in duplex for *adk* and *uidA* targets, while the amplification of *uidB* was performed in a separate reaction.

### MALDI-TOF MS

MALDI-TOF-MS spectra were obtained for the 39 Strains of *E. marmotae* and 12 other *Escherichia* isolates listed in Table 1. These spectra were also compared to MALDI-TOF-MS spectra of 176 anonymized clinical isolates that had been identified as *E. coli* in routine tests in the microbiology laboratory at Henry Ford Health. Cultures were incubated on Trypticase Soy Agar + 5% sheep blood (Remel, USA) at 35°C ± 2°C for 16-24hr. Colonies were picked and placed in a single well of a barcode-labeled VITEK MS-DS target slide (bioMérieux, France) using a VITEK Pick-Me pen and nibs (bioMérieux, France). Then the colony was covered with 1 µL of VITEK MS-CHCA Alpha-cyano-4- hydroxy-cinnamic acid (bioMérieux, France) and air-dried. All of the Henry Ford Health isolates used in this study were identified as *E. coli* by MALDI-TOF MS using the Vitek MS instrument (BioMérieux VITEK MS Software version 3.2.0) by the in vitro diagnostic (IVD) database with confidence scores of 99% and above (usually >99.9%); in rare instances, the IVD result for an *E. marmotae* strains was reported as “no identification.” We also recorded the percent confidence scores provided by Vitek’s Research Use Only (RUO) database. Images of the spectra, including the m/z values of several of the peaks identified by the Vitek software, were captured as an image for each of these isolates, as illustrated in the results.

### Statistical Analysis

The m/z values of the peaks of each strain of *E. marmotae* and *E. coli* were analyzed to compare the statistical significance of their median or average positions and ranges. Statistical analysis was performed using GraphPad Prism, employing descriptive statistics (medians, averages, variance, ranges), parametric and non-parametric and mixed methods comparisons where data were not normally distributed as determined by a Shapiro-Wilks test, and other statistical tests (e.g., outlier tests), as described with the results. Alpha levels for statistical significance were 0.05, except where Bonferroni-corrected alpha levels were used where indicated in Results.

### Whole Genome Sequencing and Data analyses

As will be described, one isolate among 176 clinical *E. coli* samples from Henry Hord Health was reclassified as a potential *E. marmotae* based on MALDI-TOF MS findings, and this identification was further confirmed by qPCR and whole genome sequencing (WGS).

For WGS, *E. marmotae* HFH1 was streaked on LB (Luria Bertani) Agar (Fisher bioreagents) and revived overnight at 37 °C. One colony was picked and grown in LB Broth (37 °C, overnight). Genomic DNA extraction utilized a DNeasy Blood & Tissue kit (Qiagen) according to the manufacturer’s instructions. Sequencing was performed on an Illumina NovaSeq 6000 Sequencer by SeqCenter in Pittsburgh, PA USA. *De novo* assembly of short reads was performed using Unicycler 0.5.0 (17). Assembly statistics were recorded with QUAST 5.2.0 (18), which measures the quality and completeness of the genome assembly. Genome annotation was performed using Prokka v1.14.6, a prokaryotic genome annotation software tool (19).

Genome analysis was performed with ABRIcate version 1.01. (https://github.com/tseeman/abricate) identified antimicrobial resistance genes using the CARD database (20), virulence genes using the *E.coli* Virulence Factor Database (21), serotype using SerotypeFinder (22) and the presence of plasmids using PlasmidFinder (23).

### Motility Assay

Bacterial motility was measured in semi-solid (0.25% agar) plates containing LB broth. The strain was grown in LB broth overnight at 37 °C with shaking at 180 rpm. Cultures were adjusted to OD_600_ = 0.06-0.1. A sterile stab was used to inoculate the culture into the center of the semi-solid agar plate (100 mm x 15 mm). Plates were incubated at 37 °C for 18-24 hours. If a bacterial spread diameter was <0.5 cm, the strain was categorized as “non-motile.”

## RESULTS

### PCR IDENTIFICATION

*In silico* analysis to compare the primers and probes in Table 2 against *E. coli* strains and other *Escherichia* genomes available in GenBank showed that the forward primer differed from all *E.coli* and other cryptic *Escherichia* clades by four bases, the reverse primer differed by two bases, and the probe differed by 3 bases, suggesting that the design would be completely specific to *E. marmotae*. Representative qPCR results illustrated in Figure 1 demonstrate the specificity of the *uidA* and *uidB* primers and probes targeting *E. marmotae*, and the efficacy of the *adk* primers and probe as a positive control amplification of all *Escherichia*. Table S1 summarizes this experimental validation as tested on 27 *E. marmotae* strains, 4 environmental *E. coli* strains, and 3 strains from other cryptic clades. Amplification of *uidA* and *uidB* was also negative on all clinical *E. coli* isolates tested (n = 175). The *uidA* and *uidB* primers and probes amplified target DNA exclusively from *E. marmotae* with 100% specificity, while the *adk* primers and probe amplified DNA from all *Escherichia* strains tested. Sanger DNA sequencing of the PCR products was done from a subset of these amplifications confirmed the correct amplification targets.

**Figure 1:**
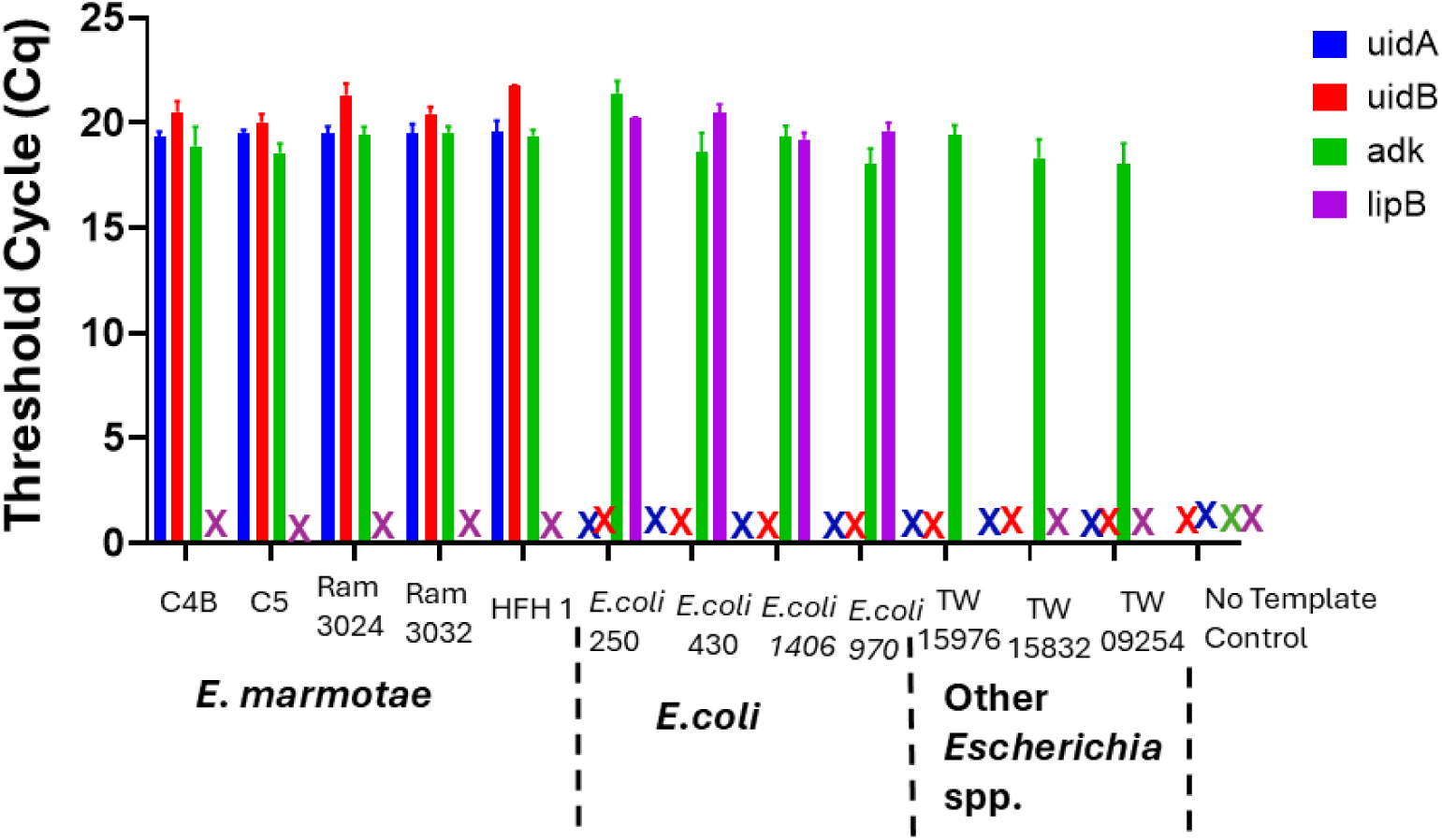
Taqman PCR identification of *E. marmotae* by species-specific primers and probes for genes *uidA* (blue),*uidB* (red), *lipB* (purple) and positive control amplification by an all *Escherichia adk-*targeted primer-probe. The “X” symbol indicates that no amplification was detected within 30 cycles. Bars show the average Cq threshold cycle number with error bars indicating the standard deviation of triplicate assays for a representative set of isolates of *E. marmotae* (C4B, C5, RAM 3024,RAM 3032 & HFH1), *E. coli*, other *Escherichia* clades and no template control.

### MALDI-TOF-MS

IVD and RUO MALDI-TOF-MS scores were determined for 176 putative *E. coli* clinical isolates from Henry Ford Health and for isolates of *E. marmotae* and other *Escherichia* listed in Table 1 that were submitted to the Henry Ford Health microbiology laboratory. The Henry Ford Health *E. coli* isolates had average IVD confidence scores of 99.9% (no variation was reported) and RUO scores in the range of 0% to 99.9% (median = 87.4%). All those that were tested (175 isolates) by our *E. marmotae/E. coli* qPCR test and an *E. coli-*specific test by (15) were confirmed as *E. coli, except one isolate which had an IVD Score of 99.1% and RUO =0%*. Similarly, as shown in Table S2, four environmental *E. coli* isolates had IVD *E. coli* confidence scores of 99.9% and RUO scores ranging from 93.9% to 99.9% (median = 98.7%). By comparison, out of 39 *E. marmotae* strains, 36 isolates had MALDI-TOF-MS IVD scores for “*E. coli*” ranging from 98.3% - 99.9% (median = 99.9%) while 3 isolates were reported as “NO ID”. The RUO scores for the 39 *E. marmotae* ranged from 0% - 84.2% (median = 0%), significantly different from the RUO scores of *E. coli* (p <0.0001; Mann-Whitney U test). Other *Escherichia* cryptic clades had IVD scores of 99.9% and RUO scores overlapping both the *E. coli* and *E. marmotae* scores (range 0% - 99.9%; median: 82.3%, p<0.0001, significantly higher than for *E. marmotae*).

Although the RUO scores of *E. marmotae* isolates indicate that significant differences exist between the spectra from *E. coli*, the RUO scores alone do not provide an absolute identification of *E. marmotae* since these scores overlap other *Escherichia* clades and even some rare variants of *E. coli*. However, by analyzing details of the mass spectra peaks provided by the “plus” version of the VITEK software, we determined that a peak between 7260 and 7268 daltons m/z was specific to *E. marmotae*; the comparable peak in *E. coli* was usually located between 7270 and 7285 daltons, and never in the range of 7260 to 7268. This is shown in representative spectra from *E. marmotae* and *E. coli*, illustrated in Figure 2, in which *E. marmotae* has a peak at m/z of 7263.837 daltons in a spectrum from a clinical sample and 7260.751 daltons in a spectrum from an environmental isolate. The comparable peak in *E. coli* has an m/z peak at approximately 7274 daltons in both environmental and clinical isolates.

**Figure 2:**
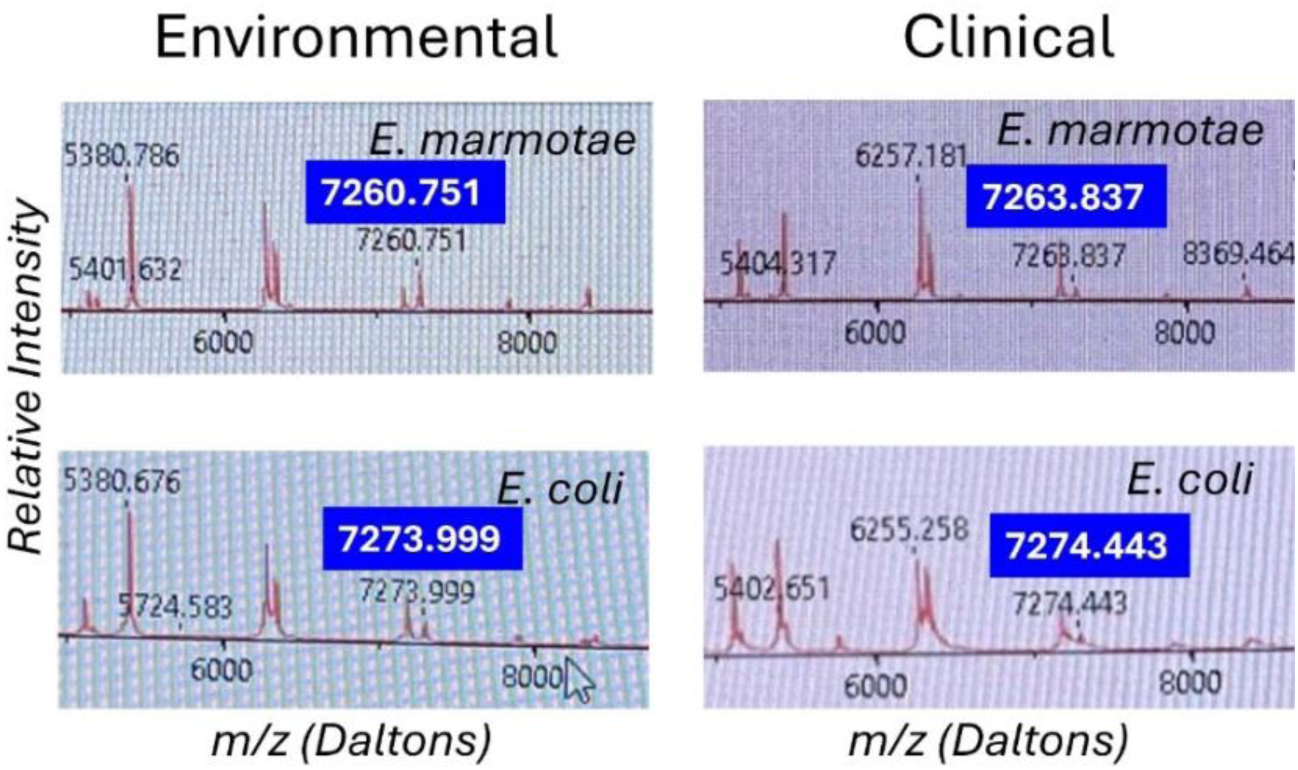
MALDI-TOF-MS spectra for *E. marmotae* and *E. coli* in the range of 5200 to 8800 daltons, highlighting the species-specific peak that usually occurs in the range of 7260 – 7268 for *E. marmotae* and between 7268 and 7285 in *E. coli*. The specific peak is highlighted in larger font for each spectrum for clarity. These representative spectra have m/z for the species-specific peak close to the median peak locations as summarized in Figure 3, for clinical and environmental strains.

Figure 3 summarizes these data statistically, comparing the m/z values both by species (*E. marmotae* vs. *E. coli*) and by whether the isolate was from a clinical or an environmental source. Since there was no overlap between *E. marmotae* and *E. coli* in the m/z values for the peak occurring in the range of 7250 – 7280 daltons, it is not surprising that the m/z for all *E. marmotae* differs significantly from all *E. coli* (unpaired t-test, p<0.0001). Environmental and clinical strains among each species had overlapping ranges for this peak within species but nevertheless differed significantly from each other for *E. marmotae* (means: environmental 7260.949; clinical 7264.137; Bonferroni-corrected unpaired t-test, p<0.0001); the *E. coli* clinical v. environmental comparison was not significant. See the supplement for additional statistical details on the comparisons in Figure 3. The statistical analysis in Figure 3 confirmed that the peak range of 7260–7268 m/z for *E. marmotae* was distinct and non-overlapping with the 7268–7285 m/z range observed for *E. coli*.

**Figure 3:**
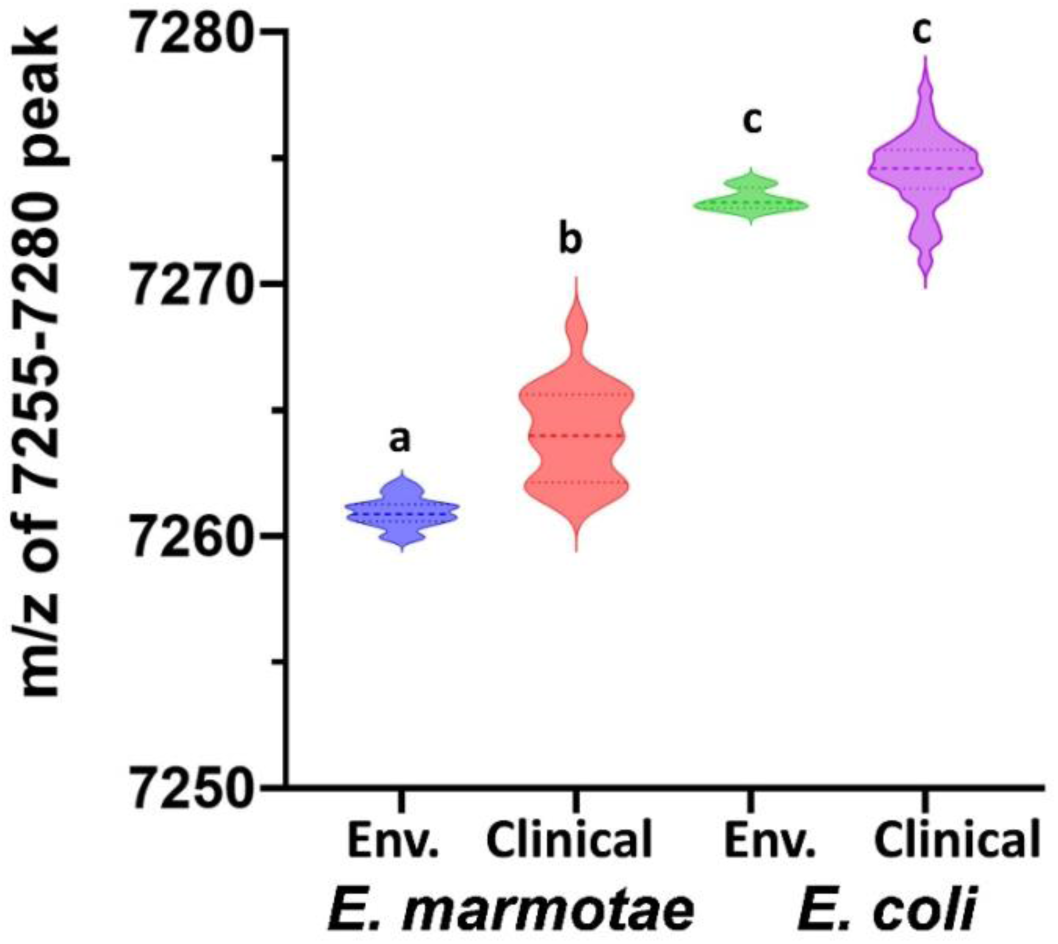
Comparative analysis of MALDI- TOF MS peaks for *E. marmotae* and *E. coli* in the m/z range of 7250 - 7280. The violin plots, with a dashed line representing the median, illustrate values of the m/z peak for 17 environmental *E. marmotae* (blue), 18 clinical *E. marmotae* (red), 4 environmental *E. coli* (green), and 169 clinical *E. coli* (purple). Groups with different letters differed from one another on statistical tests at p<0.0001 (multiple t-tests and Mann-Whitney tests, Bonferroni corrected for 6 comparisons; see supplement). The two groups with the same letter were not different (p = 0.034, not significant with the Bonferroni correction).

While most strains of *Escherichia* fit this described paradigm, a small proportion of both *E. marmotae* and *E. coli* lacked the peak in the described range, but rather had a nearby lower m/z peak. Thus, 3 of 39 *E. marmotae* isolates had a peak at 7221daltons, and one isolate had a peak at 7200 daltons. In *E. coli*, all of the environmental strains and all of the other *Escherichia* cryptic clades showed a peak in the 7270–7280 daltons m/z range (Table S1); however, among the 175 clinical *E. coli* tested from Henry Ford Health, 3 isolates had their nearby peak in the range of 7200 – 7221, as listed in Table S3 and 3 strains had a nearby peak in the range of 7280 - 7286. The idea that these low and high m/z peaks are distinctively different is supported by statistical outlier tests (GraphPad, Outlier test ROUT with Q set at 1%) on the complete data sets.

### Clinical Case Identification and Characterization of *E. marmotae* HFH1

Among the 176 clinical *Escherichia coli* isolates tested by MALDI-TOF-MS, one strain—designated HFH1—was flagged for further analysis due to its unique spectral profile. Specifically, this isolate displayed a peak at 7261.439 m/z (Figure 4), which falls within the *E. marmotae*-specific range (7260–7268 m/z) identified in our spectral analysis (Figure 2–3). Despite being identified by the IVD MALDI-TOF system as *E. coli* with a high confidence score of 99.1%, the RUO confidence score was 0%, consistent with other *E. marmotae* isolates. To further investigate, we tested the isolate with our species-specific qPCR assay targeting *E. marmotae* uidA and uidB genes. The assay had a positive result, and sequencing of the amplicons and the whole genome (HFH1 WGS assembly, completeness, and annotation statistics are given in Table S4) further confirmed its identity as *E. marmotae*. WGS of HFH1 showed that this isolate is more than 99.0% identical to other *E. marmotae* genomes in GenBank and differs from *E. coli* by 10%. The sequence of HFH1 has been submitted to GenBank as accession ID JBOZGX000000000.

**Figure 4:**
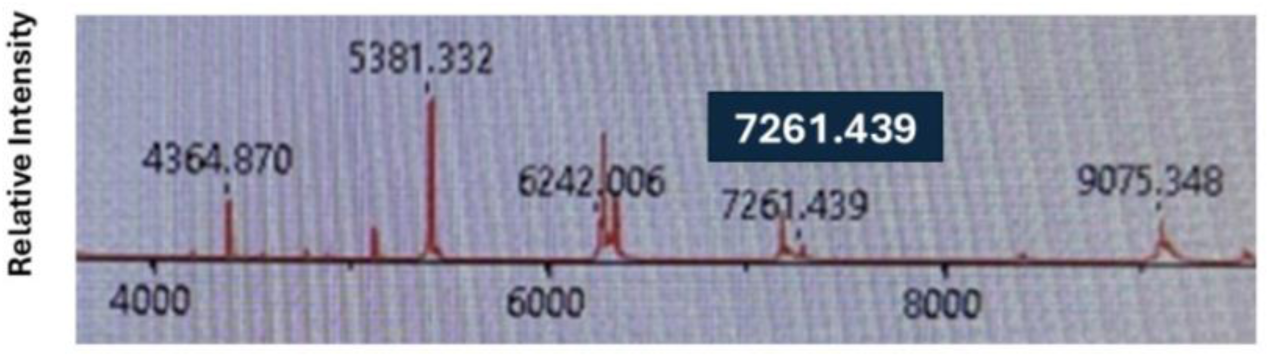
MALDI-TOF-MS spectra for *E. marmotae* HFH1 highlighting the range of *E. marmotae*-specific peak which is 7261.439. Strain HFH1 was isolated from a blood culture of a patient with clinical signs of sepsis, indicating its pathogenic potential. Motility tests revealed that HFH1 is non-motile and its serotype is O21:H56. Virulence factor profiling identified genes involved in adhesion and fimbrial assembly, such as *fimA-I, ompA, papD-H, hofB/C/Q, sfpC/H, eaeH, nada/b, fanD-H, ppdA-D*, and the curli operon *csgA/B/D/E/F/G*(24,25). The genome of HFH1 encodes genes related to flagellar biosynthesis and chemotaxis, including *flgA-N*, *flhA-E*, *fliA-Z*, *flk*, *motA/B*, and *cheA/B/R/W/Y/Z* (26). Toxin- and secretion-related genes included *cdtABC*, encoding cytolethal distending toxins that induce DNA damage in host cells (27) and *astA*, which encodes the enteroaggregative heat-stable enterotoxin EAST1. Additional virulence elements included genes for Type II (*gspC-M*), Type III (T3SS), and Type VI (T6SS) secretion systems, as well as invasion-associated loci such as *kpsD*, *kpsM*, and *ibeB/C*. The complete virulence profile is listed in Table S5 and is consistent with previously reported clinical and environmental *E. marmotae* strains (28–30)

Antimicrobial resistance (AMR) gene analysis revealed several genes capable of conferring resistance to various antibiotic classes. Genes associated with resistance to antimicrobial peptides included *eptA*, *bacA*, *ugd*, *yojI*, and *pmrF*. Nitroimidazole resistance is mediated by *msbA*, while *emrE* conferred resistance to macrolides. Multidrug efflux systems were represented by *acrA/B/E/F*, *marA*, *gadX*, *mdtE/F*, and *tolC*. Additionally, β-lactam resistance genes *ampH* and *ampC* were present, suggesting potential resistance to penams and cephalosporins. A full list of resistance genes along with sequence coverage and identity based on the CARD database in isolate HFH1 is presented in Table S6.

Plasmid analysis using PlasmidFinder revealed the presence of three plasmid replicon types in the *E. marmotae* HFH1 genome: IncX4_2, IncFIB(AP001918)_1, and IncFII(pHN7A8)_1_pHN7A8. The IncX4 replicon has been frequently associated with the global dissemination of mcr-1, a plasmid-mediated colistin resistance gene among *Enterobacteriaceae*(31,32) although no *mcr* genes were detected in HFH1. The IncFIB(AP001918) and IncFII(pHN7A8) plasmid types are commonly found in extraintestinal pathogenic *E. coli* (ExPEC) and are often linked to virulence factors and antimicrobial resistance genes (33).

The phenotypic and genotypic data confirm HFH1 as *E. marmotae* and demonstrate its potential to cause invasive human infection.

## DISCUSSION

Accuracy in identifying the organisms infecting an ill person may be a necessary pre-requisite to designing proper treatment. For an emerging pathogen, such as *E. marmotae*, which is frequently mistaken for *E. coli* because of its similar phenotype on standard tests, the lack of accurate identification means that until now the relative efficacy of different treatments for *E. marmotae* than for *E. coli* infections cannot be determined. Since the genomic sequence of *E. marmotae* differs on average from *E coli* by about 10%, sequence-based tests can easily be applied; however, to our knowledge, no FDA-approved PCR test to do so is commercially available. We have demonstrated here two Taqman PCR targets that specifically identify *E. marmotae* and could be used in such tests.

MALDI-TOF-MS is a frequently used clinical laboratory method for fast identification of bacterial isolates; however, MALDI-TOF-MS databases do not universally include *E. marmotae*. The Bruker reference database includes several *E. marmotae* spectra, but publications reported that the confidence scores with the current database are relatively low (6,7). The Biomérieux VITEK MALDI-TOF-MS database does not include *E. marmotae* at all, and therefore, as reported here, either mistakenly identifies verified *E. marmotae* as *E. coli* most of the time or doesn’t identify the isolate at all.

However, we report here a specific spectral peak observed on the VITEK MALDI-TOF-MS that unambiguously identifies most strains of *E. marmotae*, differentiating them, with no overlap, from isolates of *E. coli.* The species-specific spectral peak in *E. marmotae* occurs at m/z of 7260 – 7268, while the comparable peak in *E. coli* is at 7268 – 7286. A small percentage (<10%) of both species lacks the peak at these m/z values, but seem to have a peak that is otherwise absent in the m/z range of 7200 – 7225. If the isolate has a peak in the range of 7260 – 7268, this study unambiguously identifies it as *E. marmotae*.

Until now, the prevalence of *E. marmotae* in clinical cases appears to be low compared to *E. coli* and have previously only been reported in publications from Norway, France, and Australia (5–7); This study provides the first documented clinical identification of *E. marmotae* in the United States, highlighting the need for updated diagnostic protocols capable of detecting this pathogen.

The prevalence of *E. marmotae* compared to *E. coli* is below 1% (0.2% in France (5); and 0.4% in Norway (7). In the present study, in which almost 200 clinical isolates putatively identified as *E. coli* were tested for being *E. marmotae,* one strain, HFH1, proved to be *E. marmotae*; the rest were confirmed as *E. coli*. With only one isolate out of 176, we cannot calculate an accurate prevalence, but this frequency is consistent with a prevalence of less than 1%, as reported elsewhere.

The isolate was recovered from a patient with sepsis, indicating its potential to cause invasive disease. Motility analysis revealed that HFH1 is not motile at 37 °C and therefore may cause a different range of infections than *E. coli*, which is typically motile at this temperature. The temperature dependence of motility and motility gene expression of *E. marmotae*, in comparison to *E. coli*, is the subject of another study from our laboratory (11). Genetic analysis of HFH1 also reveals numerous non-synonymous sequence differences from *E. coli* in genes that are important for extraintestinal pathogenic *E. coli* (ExPEC) infections, including genes for fimbrial adhesion, toxin production, and multiple secretion systems.

*E. marmotae* is known to occur in the environment in the USA, where it was originally identified as a “cryptic clade” of *Escherichia* (2). Although the identifying MALDI TOF MS peak in the environmental strains of *E marmotae* in the present study occurs at a lower m/z than most clinical strains we have tested, our HFH1 isolate is closer to the average environmental isolate (all of which were from Michigan) than the average clinical isolate of *E. marmotae*, most of which were from Norway. The significant difference may therefore be one of geographical origin and not environment vs. clinic. Future studies may investigate these differences in the identifying peak.

Clinical tests based on these observations may assist in therapeutic decisions. Moreover, discovering the identity of the structure underlying the shift in the MALDI-TOF-MS peak may help determine if it has a special role in infection; however, until now the molecule(s) that is detected by this peak are unknown. Although currently rare among clinical isolates of *Escherichia*, it seems likely that additional clinical isolates will be detected globally. We wonder whether *E. marmotae* is an emerging pathogen or whether it has always been rare and will remain so. Future research should explore the host immune response to *E. marmotae*, the functional role of its unique proteomic features, and comparative infection models to elucidate how its pathogenesis diverges from that of *E. coli*.

## Data Availability

All data produced in the present work are contained in the manuscript

## Abbreviations

MALDI-TOF MS: Matrix-Assisted Laser Desorption Ionization-Time of Flight Mass Spectrometry
IVD: in vitro diagnostic
RUO: Research Use Only
AMR: Antimicrobial resistance
LB: Luria Bertani
WGS: Whole Genome Sequencing
CARD: Comprehensive Antibiotic Resistance Database
ExPEC: Extraintestinal Pathogenic *Escherichia coli*
FDA: Food and Drug Administration
qPCR: Quantitative Polymerase Chain Reaction
PCR: Polymerase Chain Reaction
m/z: Mass-to-charge ratio
T3SS: Type III Secretion System
T6SS: Type VI Secretion System
IDT: Integrated DNA Technologies

## DECLARATIONS

### Ethics approval and consent to participate

The *E. coli* and *E. marmotae* isolates used in this research were collected from Henry Ford Health. All patient data were anonymized, prior to research use.

*E. marmotae* cultures from Norway were collected in a study approved by the Regional Ethical Committee of Western Norway (REK 322324) (7). This research adhered to the Declaration of Helsinki.

### Availability of data and materials

The raw sequencing data and genome assemblies of the RAM lab environmental and clinical isolates have been published under the BioProject PRJNA1261436 and PRJNA1268432, and accession numbers are provided in Table 1.

### Consent for publication

All authors contributed to writing – review & editing and approved the final version of the manuscript.

### Authors’ contributions

P.M.O. conceived the study and led conceptualization, data curation, investigation, software, formal analysis, and writing – original draft. R.J.T., J.M.B., M.S., and M.Z. contributed resources, methodology and supported data acquisition from Henry Ford Health. A.S. and T.S.B. contributed to data validation and investigation. A.F. supported investigation and laboratory analyses. J.L.R. contributed to data curation, formal analysis, methodology, project administration, software, resources, writing—editing of original draft, supervision and validation.

## Acknowledgements

Not Applicable

