## Supplemental Table 1-4 for "New Diagnostic Methods for *Escherichia marmotae* and the First Report of its Identification in Clinical Isolates in North America"

Table of Contents.

**Table S1: Average Cq threshold cycle number of strains of *E. marmotae*, *E. coli* and *Escherichia* cryptic clades**

**Table S2: MALDI SPECTRA result showing IVD and RUO scores and the specific peak in *E. marmotae,* *E.coli* and other *Escherichia* cryptic clades.**

**Supplementary Data:**

**Table S1: Average Cq threshold cycle number of strains of *E. marmotae*, *E. coli* and *Escherichia* cryptic clades**

| Strains | *uidA* | *uidB* | *adk* | *lipB* |
| --- | --- | --- | --- | --- |
| C4B | 19.33 | 20.51 | 18.87955 | ND |
| C5 | 19.51 | 20.01 | 18.5445 | ND |
| C6 | 19.14 | 20.41 | 19.10844 | ND |
| C1 | 19.46 | 20.45 | 18.23005 | ND |
| C7 | 20.09 | 20.18 | 18.5719 | ND |
| C8 | 19.17 | 19.89 | 19.68103 | ND |
| AS8 | 19.76 | 20.34 | 20.34157 | ND |
| C9 | 19.69 | 20.06 | 18.39248 | ND |
| C10 | 19.26 | 20.57 | 18.28248 | ND |
| C11 | 19.96 | 20.29 | 19.34203 | ND |
| C12 | 19.61 | 20.73 | 18.34626 | ND |
| C3 | 19.51 | 20.45 | 19.43225 | ND |
| AS14 | 19.79 | 20.27 | 18.30907 | ND |
| AS15 | 19.78 | 20.50 | 18.34307 | ND |
| AS16 | 19.97 | 20.47 | 18.91359 | ND |
| AS17 | 19.56 | 20.65 | 19.74667 | ND |
| AS18 | 19.78 | 20.30 | 18.33043 | ND |
| C2 | 20.07 | 20.26 | 19.37026 | ND |
| C4A | 19.76 | 20.14 | 18.79661 | ND |
| AS21 | 20.15 | 20.56 | 18.56578 | ND |
| AS22 | 19.59 | 20.84 | 19.28115 | ND |
| AS23 | 20.55 | 20.61 | 18.9066 | ND |
| RAM 3024 | 19.51 | 21.31 | 19.42667 | ND |
| RAM 3032 | 19.47 | 20.42 | 19.53464 | ND |
| RAM 3054 | 17.92 | 21.07 | 18.39333 | ND |
| RAM 3050 | 19.08 | 21.90 | 19.11333 | ND |
| TW 09308 | 19.44 | 20.99 | 19.30333 | ND |
| RAM HFH1 | 19.71 | 21.72 | 19.35 | ND |
| E.coli 250 | ND | ND | 21.39044 | 20.18 |
| E.coli 430 | ND | ND | 18.61581 | 20.49 |
| E.coli 1406 | ND | ND | 19.34203 | 19.41 |
| E.coli 970 | ND | ND | 18.01292 | 19.60 |
| TW 15976 | ND | ND | 19.43225 | ND |
| TW 15832 | ND | ND | 18.30907 | ND |
| TW 09254 | ND | ND | 18.00973 | ND |

**Table S2: MALDI SPECTRA result showing IVD and RUO scores and the specific peak in *E. marmotae,* *E.coli* and other *Escherichia* cryptic clades.**

| **Strains** | **IVD score** | **RUO scores** | **Specie Specific Peak** |
| --- | --- | --- | --- |
| TW14263 alias Ram 3024 | 98.3 | 0 | 7261.267 |
| TW14264 alias Ram 3032 | 99.9 | 0 | 7261.267 |
| TW14265 alias Ram 3050 | 99.9 | 0 | 7260.376 |
| TW14266 alias Ram 3054 | 99.9 | 0 | 7260.878 |
| TW14267 alias Ram 3318 | 99.9 | 77.6 | 7261.133 |
| TW 09308 | 99.9 | 0 | 7261.649 |
| TW 14351 | 99.9 | 0 | 7260 |
| TW 15825 | 99.9 | 0 | 7261.82 |
| TW 15841 | 99.9 | 75.2 | 7259.911 |
| TW 15835 | 99.9 | 84.2 | 7260.802 |
| TW 15848 | 99.9 | 0 | 7260.802 |
| TW 15839 | 99.9 | 0 | 7261.184 |
| TW 15833 | 99.9 | 0 | 7262.075 |
| TW 15846 | 99.9 | 0 | 7261.148 |
| TW 15836 | 99.9 | 0 | 7260.639 |
| TW 15840 | 99.9 | 0 | 7260.511 |
| TW 15834 | 99.9 | 0 | 7260.674 |
| C4B | 99.9 | 0 | 7263.837 |
| C5 | No ID | 0 | 7221.083 |
| C6 | 99.9 | 0 | 7263.964 |
| C1 | NO ID | 0 | 7200.514 |
| C7 | 99.9 | 0 | 7265.111 |
| C8 | 99.9 | 0 | 7264.000 |
| AS8 | NO ID | 0 | 7265.493 |
| C9 | 99.9 | 0 | 7265.621 |
| C10 | 99.9 | 0 | 7265.621 |
| C11 | 99.9 | 0 | 7264.729 |
| C12 | 99.9 | 0 | 7268.298 |
| C3 | 99.9 | 0 | 7266.131 |
| AS14 | 99.9 | 0 | 7201.148 |
| AS15 | 99.9 | 0 | 7201.148 |
| AS16 | 99.9 | 0 | 7266.131 |
| AS17 | 99.9 | 0 | 7262.128 |
| AS18 | 99.9 | 0 | 7261.364 |
| C2 | 99.9 | 0 | 7261.746 |
| C4A | 99.9 | 0 | 7262.128 |
| AS21 | 99.9 | 0 | 7262.765 |
| AS22 | 99.9 | 0 | 7261.874 |
| AS23 | 99.9 | 0 | 7263.529 |
| TW 15976 | 99.9 | 80.0 | 7274.891 |
| TW 15838 | 99.9 | 83.4 | 7274.008 |
| TW 15951 | 99.9 | 99.9 | 7274.381 |
| TW 15832 | 99.9 | 0.00 | 7274.008 |
| TW 09254 | 99.9 | 87.2 | 7273.361 |
| TW 09231 | 99.9 | 87.6 | 7274.253 |
| TW 14182 | 99.9 | 81.2 | 7273.999 |
| TW 11588 | 99.9 | 80.0 | 7273.498 |
| E.coli 250 | 99.9 | 99.0 | 7272.988 |
| E.coli 430 | 99.9 | 99.9 | 7273.370 |
| E.coli 1406 | 99.9 | 98.4 | 7273.106 |
| E.coli 970 | 99.9 | 93.9 | 7273.999 |

**Table S3: MALDI SPECTRA result showing IVD and RUO scores and the specific peak in *clinical*  *E.coli* from Henry Ford Laboratory**

| Strains | IVD Scores | RUO scores | Peaks |
| --- | --- | --- | --- |
| M43600-1 | 99.9 | 0 | 7218.072 |
| T501076 - 1 | 99.9 | 98.5 | 7270.61 |
| S382911 - 1 | 99.9 | 0 | 7270.816 |
| H594473 - 1 | 99.9 | 0 | 7270.865 |
| S424820 - 1 | 99.9 | 93.5 | 7271.049 |
| X141675 - 2 | 99.9 | 81.5 | 7271.071 |
| F466878 - 1 | 99.9 | 82.5 | 7271.584 |
| W450286-1 | 99.9 | 0 | 7271.601 |
| F434517 - 1 | 99.9 | 90 | 7271.651 |
| M446123 - 1 | 99.9 | 87.5 | 7271.703 |
| H2785 - 1 | 99.9 | 83.3 | 7271.736 |
| W58342 - 1 | 99.9 | 87.2 | 7271.819 |
| F516168 - 1 | 99.9 | 93.8 | 7271.827 |
| X102872 - 1 | 99.9 | 97.8 | 7271.911 |
| T579217 - 1 | 99.9 | 95.2 | 7271.937 |
| H587114 - 2 | 99.9 | 87.3 | 7271.994 |
| T362091-1 | 99.9 | 0 | 7272.026 |
| T483117 | 99.9 | 95.2 | 7272.219 |
| W75726 - 1 | 99.9 | 82.5 | 7272.223 |
| X141675 -1 | 99.9 | 0 | 7272.344 |
| M583411 - 1 | 99.9 | 75.4 | 7272.347 |
| M568608 - 1 | 99.9 | 99.9 | 7272.394 |
| W682619-1 | 99.9 | 77.5 | 7272.42 |
| F351529 - 1 | 99.9 | 89.3 | 7272.745 |
| M377597 - 1 | 99.9 | 86.4 | 7272.901 |
| T261647-1 | 99.9 | 98.4 | 7273.117 |
| X269308-2 | 99.9 | 99.9 | 7273.27 |
| W371120 - 2 | 99.9 | 87.6 | 7273.323 |
| S518994 - 1 | 99.9 | 96.3 | 7273.345 |
| M588863 - 1 | 99.9 | 86.5 | 7273.402 |
| M471433 - 4 | 99.9 | 82.5 | 7273.435 |
| X210193 - 2 | 99.9 | 99.9 | 7273.441 |
| T370915-1 | 99.9 | 87.5 | 7273.518 |
| W41195 - 1 | 99.9 | 86.3 | 7273.544 |
| M542834 - 3 | 99.9 | 0 | 7273.551 |
| H699340 - 1 | 99.9 | 99.9 | 7273.591 |
| M41468-1 | 99.9 | 90.5 | 7273.754 |
| X283079-1 | 99.9 | 99.9 | 7273.758 |
| T94348 - 2 | 99.9 | 99.9 | 7273.829 |
| W689970-1 | 99.9 | 0 | 7273.876 |
| W140653 - 1 | 99.9 | 83.3 | 7273.897 |
| T286241 | 99.9 | 87.2 | 7273.915 |
| H449992-1 | 99.9 | 85 | 7273.962 |
| M62714 | 99.9 | 87.5 | 7273.98 |
| H88796 - 1 | 99.9 | 75.8 | 7273.991 |
| F704616-1 | 99.9 | 90.2 | 7273.994 |
| H567166-1 | 99.9 | 99.9 | 7273.638 |
| H570493-1 | 99.9 | 79.6 | 7275.210 |
| H566692-1 | 99.9 | 93.1 | 7274.405 |
| H566692-2 | 99.9 | 90.2 | 7273.406 |
| H565851-1 | 99.9 | 0 | 7272.648 |
| T482874-1 | 99.9 | 98.4 | 7273.384 |
| H577637-1 | 99.9 | 97.5 | 7272.963 |
| H577637-2 | 99.9 | 99.9 | 7275.343 |
| F270595-1 | 99.9 | 96.7 | 7274.282 |
| H573512-1 | 99.9 | 86.5 | 7274.647 |
| S518900 - 1 | 99.9 | 0 | 7274.009 |
| M61647-1 | 99.9 | 95.4 | 7274.027 |
| S520458 - 1 | 99.9 | 0 | 7274.074 |
| F544368 - 2 | 99.9 | 99.9 | 7274.101 |
| W61945 - 1 | 99.9 | 92.3 | 7274.11 |
| W156575 - 4 | 99.9 | 98.4 | 7274.166 |
| W686297-1 | 99.9 | 83.3 | 7274.176 |
| W674048-3 | 99.9 | 80.1 | 7274.189 |
| H565879-2 | 99.9 | 99.9 | 7274.2 |
| W570445-1 | 99.9 | 92.4 | 7274.232 |
| W443456-2 | 99.9 | 95.5 | 7274.25 |
| W560015-2 | 99.9 | 86.3 | 7274.256 |
| W558193-1 | 99.9 | 84.7 | 7274.283 |
| X688583-1 | 99.9 | 97.6 | 7274.295 |
| H75885 - 1 | 99.9 | 83.4 | 7274.319 |
| T374094-2 | 99.9 | 89.3 | 7274.358 |
| W425622-2 | 99.9 | 0 | 7274.359 |
| F373521 - 3 | 99.9 | 0 | 7274.366 |
| M479312 - 1 | 99.9 | 75.4 | 7274.39 |
| X285519-1 | 99.9 | 85.1 | 7274.396 |
| T629234 - 1 | 99.9 | 93.1 | 7274.419 |
| W164580 - 1 | 99.9 | 85.4 | 7274.443 |
| T485744 | 99.9 | 92.9 | 7274.447 |
| H16219 - 1 | 99.9 | 82.5 | 7274.448 |
| H109142 - 2 | 99.9 | 0 | 7274.456 |
| X138248 - 11 | 99.9 | 83 | 7274.457 |
| M149181-2? | 99.9 | 87.2 | 7274.475 |
| M577899 - 1 | 99.9 | 81.4 | 7274.505 |
| H688852 - 1 | 99.9 | 81.4 | 7274.512 |
| X185929 - 1 | 99.9 | 77.9 | 7274.536 |
| M13245-2 | 99.9 | 99.9 | 7274.544 |
| T361221-2 | 99.9 | 87.2 | 7274.555 |
| M164781-1 | 99.9 | 93 | 7274.591 |
| T260545-1 | 99.9 | 81.4 | 7274.677 |
| F508018 - 1 | 99.9 | 93.5 | 7274.695 |
| M414328 - 1 | 99.9 | 93.1 | 7274.703 |
| W371126 - 1 | 99.9 | 99.9 | 7274.725 |
| M259498-2 | 99.9 | 87.2 | 7274.727 |
| W175456 - 1 | 99.9 | 77.6 | 7274.749 |
| F684244-1 | 99.9 | 90.1 | 7274.763 |
| W138610 - 1 | 99.9 | 75.8 | 7274.777 |
| T491542-1 | 99.9 | 91.9 | 7274.786 |
| T260545-2 | 99.9 | 87.2 | 7274.805 |
| F710862-1 | 99.9 | 87.5 | 7274.806 |
| X155929 - 2 | 99.9 | 99.9 | 7274.812 |
| W174094 - 1 | 99.9 | 83.9 | 7274.825 |
| M164781-2 | 99.9 | 99.9 | 7274.846 |
| M259498-3 | 99.9 | 97.7 | 7274.854 |
| W202658 - 1 | 99.9 | 83.3 | 7274.855 |
| H685711 - 1 | 99.9 | 93.1 | 7274.922 |
| W32749 - 1 | 99.9 | 0 | 7274.951 |
| T273544-1 | 99.9 | 82.4 | 7274.969 |
| H449992-2 | 99.9 | 89.4 | 7274.982 |
| X106451 - 1 | 99.9 | 98.9 | 7274.997 |
| T66906 - 1 | 99.9 | 99.9 | 7275.057 |
| W434641-1 | 99.9 | 99.9 | 7275.09 |
| T620280 - 1 | 99.9 | 84.3 | 7275.093 |
| H568852-1 | 99.9 | 86.4 | 7275.1 |
| F267089-1 | 99.9 | 0 | 7275.108 |
| M381064 - 1 | 99.9 | 87.2 | 7275.162 |
| M506012 - 1 | 99.9 | 90 | 7275.171 |
| W82552 - 1 | 99.9 | 97.5 | 7275.184 |
| S502577 - 3 | 99.9 | 98.5 | 7275.185 |
| W681466 | 99.9 | 84.4 | 7275.209 |
| W463421 | 99.9 | 99.9 | 7275.215 |
| M403605 -2 | 99.9 | 99.9 | 7275.218 |
| M155733-2 | 99.9 | 80.8 | 7275.223 |
| H660901 - 1 | 99.9 | 99.9 | 7275.234 |
| H565879-1 | 99.9 | 99.9 | 7275.276 |
| W443191-1 | 99.9 | 98.4 | 7275.302 |
| X688015-9 | 99.9 | 90 | 7275.303 |
| T356776-3 | 99.9 | 87.5 | 7275.322 |
| S514130 - 1 | 99.9 | 81.3 | 7275.327 |
| W460868-2 | 99.9 | 99.9 | 7275.333 |
| W384925 - 1 | 99.9 | 93.7 | 7275.356 |
| M362604 - 1 | 99.9 | 99.9 | 7275.448 |
| W683804 | 99.9 | 83.7 | 7275.464 |
| W595366-1 | 99.9 | 79.3 | 7275.469 |
| H146023 - 2 | 99.9 | 99.9 | 7275.515 |
| H263911 - 1 | 99.9 | 80.3 | 7275.553 |
| S397665 - 1 | 99.9 | 83.2 | 7275.576 |
| M546434 - 1 | 99.9 | 0 | 7275.587 |
| W681465 | 99.9 | 78.4 | 7275.591 |
| X215180 - 1 | 99.9 | 90.9 | 7275.608 |
| W450382-1 | 99.9 | 0 | 7275.679 |
| T581445 - 1 | 99.9 | 95.3 | 7275.703 |
| T356776-4 | 99.9 | 86.4 | 7275.705 |
| T367996-1 | 99.9 | 0 | 7275.727 |
| H660901 - 2 | 99.9 | 80 | 7275.728 |
| W200381 - 1 | 99.9 | 99.9 | 7275.743 |
| H650703 - 2 | 99.9 | 88.6 | 7275.831 |
| T497129 | 99.9 | 87.2 | 7275.915 |
| T620280 - 6 | 99.9 | 92.4 | 7275.923 |
| T674672 - 1 | 99.9 | 84.3 | 7275.924 |
| M507603 - 1 | 99.9 | 95.4 | 7275.989 |
| T484107 | 99.9 | 84.5 | 7276.08 |
| T361224-1 | 99.9 | 0 | 7276.084 |
| M152234-1 | 99.9 | 0 | 7276.096 |
| T274016-1 | 99.9 | 89.3 | 7276.11 |
| F518804 - 1 | 99.9 | 99.9 | 7276.346 |
| H29738 - 1 | 99.9 | 89.4 | 7276.394 |
| T696469 - 1 | 99.9 | 84.1 | 7276.44 |
| H650703 - 1 | 99.9 | 84.6 | 7276.468 |
| F334315 - 2 | 99.9 | 78 | 7276.637 |
| H650703 - 3 | 99.9 | 90.5 | 7276.851 |
| W682619-2 | 99.9 | 0 | 7276.878 |
| S452562 - 1 | 99.9 | 95.3 | 7276.957 |
| M365126 - 2 | 99.9 | 95.4 | 7277.175 |
| F640317 - 1 | 99.9 | 85.1 | 7277.244 |
| M503941 - 1 | 99.9 | 99.9 | 7277.652 |
| T303591-2 | 99.9 | 99.9 | 7277.669 |
| S389864 - 1 | 99.9 | 87 | 7277.673 |
| M150120-1 | 99.9 | 99.9 | 7277.881 |
| M60781-1 | 99.9 | 92.1 | 7278.279 |
| H263748 - 1 | 99.9 | 77.5 | 7281.616 |
| M152848-1 | 99.9 | 0 | 7281.96 |
| H687065 - 2 | 99.9 | 0 | 7285.062 |
| W681467 | 99.9 | 0 | 7215.946 |
| M325108-1 | 99.9 | 0 | 7217.761 |
| RAM HFH1 | 99.1 | 0 | 7261.439 |

Table S4: Summary of Whole Genome Sequencing Assembly, Completeness and Gene Annotation of *E. marmotae* Clinical Isolate HFH1

| Parameters | Data |
| --- | --- |
| Assembly | |
| Number of Contigs | 74 |
| Total length (bp) | 4614133 |
| GC Content (%) | 50.37 |
| N50 | 177349 |
| N75 | 96904 |
| L50 | 8 |
| L75 | 16 |
| Annotation of the Genome | |
| Total number of Genes | 4353 |

TABLE S5: Complete Virulence Genes list of *E. marmotae* HFH1. See supplemental Excel file.

TABLE S6: Antibiotic Resistance Genes oF *E. marmotae* HFH1. See supplemental Excel file.
